# Development of an open-source artificial intelligence (AI) anonymizer for electrocardiogram scans

**DOI:** 10.1101/2025.10.19.25338291

**Authors:** Gary Tse, Haipeng Liu, Angelina Amabi Perez Aranda, Wing Tak Wong, Sharen Lee, Tong Liu, Vellaisamy A L Roy, Branislav Milovanovic, Mehrdad Shahmohammadi Beni

## Abstract

The application of artificial intelligence (AI) in the medical field has seen a significant increase in popularity, particularly for its ability to accurately detect abnormalities across a range of diagnostic tests. The effectiveness and precision of AI models are highly contingent on the quality and diversity of the training data used in their development. In the present work, we have developed an open-source AI model designed to anonymize electrocardiogram (ECG) recordings. This model achieves anonymization by automatically detecting and extracting the waveform data. This tool can be used to prepare input data that in turn serve as input variables for training AI models specifically for cardiology applications. By ensuring that patient-identifying information is removed while retaining the essential waveform data. The present model facilitates the creation of robust, privacy-preserving datasets that can enhance the training and performance of AI in cardiology.

## 1 Introduction

Artificial intelligence (AI), including machine learning, models in medical applications are increasingly common in biomedical research; this is to train numerical models for the detection of abnormalities in different investigations. Various forms of data may contain identifiable personal information [1] and in turn pose serious privacy concerns.

Electrocardiography (ECG) is an important and widely used non-invasive method for screening and diagnosing cardiac and non-cardiac conditions. The recorded ECG contains patient-specific information such as name, age, identification number, sex, clinical indication and other demographics, these details are typically included as part of the ECG report [2] [3] [4].

Across the literature, a shared theme is the growing importance of digital and AI-based methods to enhance electrocardiogram (ECG) analysis and clinical utility. Fully automated digitization tools and deep learning models have demonstrated high fidelity, enabling the transformation of legacy ECG archives into reusable datasets for AI training [5]. Likewise, another framework employs effective preprocessing (e.g., shadow removal) before passing digitized signals to deep learning algorithms, achieving 97% binarization accuracy and 94.4% accuracy for ECG-based disease classification [6]. AI-Driven ECG Interpretation and Risk Prediction Several reviews underscore how AI—especially deep learning—has effectively reached, or even surpassed, human-level diagnostic accuracy across diverse cardiovascular conditions [7] [8] [9] [10]. Complex models such as convolutional neural networks (CNNs) can recognize subclinical patterns indicative of left ventricular dysfunction, episodic atrial fibrillation, valvular disease, hypertrophic cardiomyopathy, and even systemic illnesses (e.g., COVID-19) from standard ECGs [7] [8]. Emerging evidence also highlights that AI-ECG tools can predict demographic features (e.g., age, sex, race) and immunologic status, improving both precision medicine and public health screening [9]. These studies demonstrate a growing reliance on massive ECG datasets to fuel AI-driven insights, which underscores the critical need for robust privacy measures and also a practical solution.

In another study, the limitations of thermal paper recordings was highlighted and a simple method using scanning 12-lead ECG paper was proposed and the scanned images was then processed based on their grid sizes and signals were extracted [11]. The proposed process was simply an image segmentation process that was used for partitioning of an ECG as a digital image into multiple segments (sets of pixels), so that features in each segment could be analyzed. The proposed method of extracting ECG signals from scanned thermal paper inherently relies on the visibility of the printed grid lines, which serve as a reference for segmentation. Consequently, if the grid is faint or entirely absent—due to poor print quality, fading, or other artifacts—this approach will be prone to errors or may fail altogether. Moreover, scanning numerous ECG sheets is labor-intensive and time-consuming. Each sheet must be loaded into the scanner, and the quality of the scanned image (including the visibility of crucial grid lines) cannot be confirmed until after the scanning is completed. This makes it difficult to address issues such as misalignment, poor resolution, or faint grids in real time, further complicating the reliable extraction of ECG signals. Considering our present model which is an open-source anonymization method that could help detect and extract ECG waveforms either using scanned images, smartphone camera pictures and video feeds for batch detection, would be useful in these scenarios and the model is both timely and complementary, potentially filling a gap not fully explored in the present literature.

Previously, the digitization of ECG scans has been achieved with the help of machine learning [4] [12] by employing enormous number of ECG scans for the training of the model. Considering this, it would be pertinent to fully anonymize these ECG scans that will be used in training and digitization of machine learning models, which in turn will reduce the privacy concerns and enhance the ease of training. In present work, we have trained and developed an open-source AI model with an easy-to-use interface for anonymizing ECG results. Extracting ECG waveforms from visual formats such as images, video clips, or live feeds is often a manual and labor-intensive task, requiring additional preprocessing steps and specialized software tools. Our AI model automates this process by detecting and cropping ECG waveforms directly from multimedia inputs, simplifying data preprocessing while ensuring clean and isolated waveforms for further analysis. This model has diverse applications, including automatic cropping of ECG waveforms for efficient digitization, creating clean visuals for educational materials, supporting archival and retrieval of older ECG data, enabling real-time waveform extraction from live video feeds in emergency or remote monitoring settings, and providing highquality input data for AI-based ECG interpretation. The key advantages of our model include eliminating manual effort, reducing processing time and errors, ensuring anonymization of sensitive patient data, and supporting various media formats, making it adaptable across different environments. As an open-source solution, it democratizes access to ECG digitization tools previously confined to specialized domains or proprietary software.

## 2 Methodology

### 2.1 Training AI model based on Yolov5

In the present work, we have used *∼*3700 ECG data from patients admitted to a single tertiary hospital from Hong Kong, with variety of image qualities, shadings, grid colors and orientations. This study arose directly from clinical data collected as part of GT’s doctoral thesis at the Faculty of Medicine, Division of Graduate Studies, The Chinese University of Hong Kong accompanied by an ethics approval from the Joint NTEC-CUHK Clinical Research Ethics Committee (CUHK-NTEC CREC 2019.338 and 2019.361). In addition, the label containing patient-specific information was found to be at various locations on the ECG scans. In present work, we mainly focused on extracting the waveform section from the entire ECG scan that contains patient-specific information. Therefore, we have labelled the ECG scans using one class named “waveform” using labelImg (https://pypi.org/project/labelImg/) tool. The position of the waveform class was selected using a box and its coordinate relative to the ECG scan was saved in XML file for each ECG scan. These data were then used to train our AI model based on the widely used YOLO (You Only Look Once) [13] state-of-the-art, real-time object detection system version 5 which is based on Yolov1-Yolov4 [14]. The entire backend part of the model for AI training was written in Python programming language. The trained model was used in the inference part of the present work to separate the waveform features from other information on the ECG scans.

The flowchart of the inference model is shown in Fig. 1.

**Figure 1.**
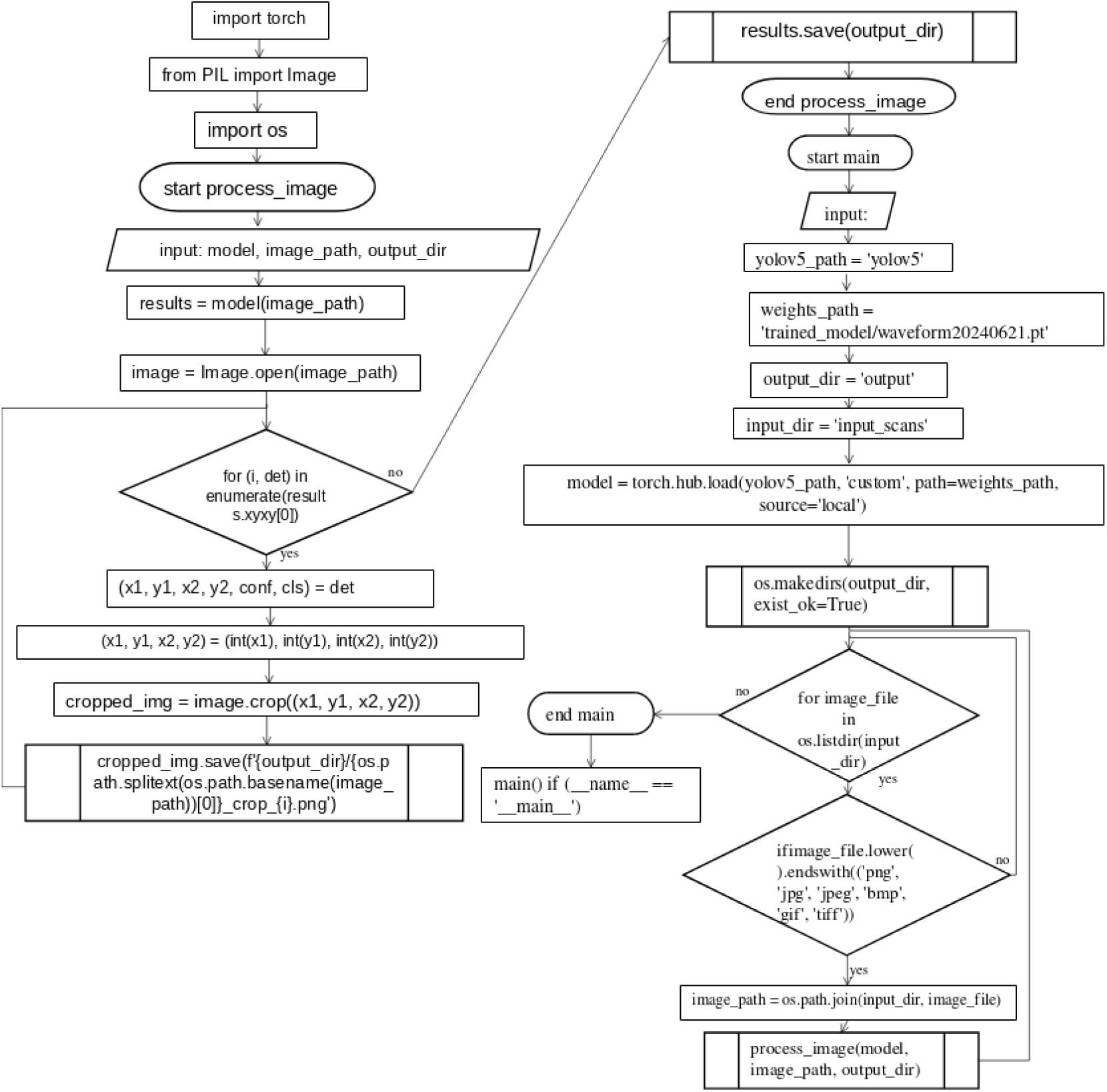
The flowchart of the ECG waveform inference module.

Training object detection models like YOLOv5, YOLOv9, and Fast R-CNN that were employed in the present work rely on supervised learning, where labeled data plays a crucial role. The process begins with dataset preparation, which involves collecting images containing waveforms and annotating bounding boxes that highlight their positions. These annotations, often stored in formats like COCO or Pascal VOC, define the ground truth for the model to learn from. Pre-trained weights, often obtained from training on large-scale datasets like COCO, serve as the starting point for fine-tuning on the waveform dataset. The training involves optimizing the model’s parameters using algorithms like Stochastic Gradient Descent (SGD) or Adam, minimizing a loss function that combines localization and classification errors.

YOLOv5 and YOLOv9 employ advanced architectures based on convolutional neural networks (CNNs), featuring a single-stage detection pipeline for real-time performance. These models integrate techniques like CSPNet and path aggregation for efficient feature extraction and merging, allowing them to perform well on diverse datasets, including waveform images. Fast R-CNN, on the other hand, follows a two-stage approach, first generating region proposals and then classifying and refining them. This approach provides higher accuracy at the cost of speed. During training, data augmentation techniques such as flipping, rotation, and random cropping enhance model generalization. Transfer learning accelerates convergence, while hyperparameter tuning optimizes model performance for waveform detection. These models benefit from frameworks like PyTorch or TensorFlow, which facilitate efficient model implementation and scaling.

### 2.2 Developed inference models

Total of five different model variations were built using our trained model, namely, (1) docker images, (2) command line interface (CLI), (3) movie analyzer, (4) live video feed, (5) web-based flask application. These developed inference models were shown schematically in Fig. 2; these utilize the trained AI model to perform inference on the user input ECG scans.

**Figure 2.**
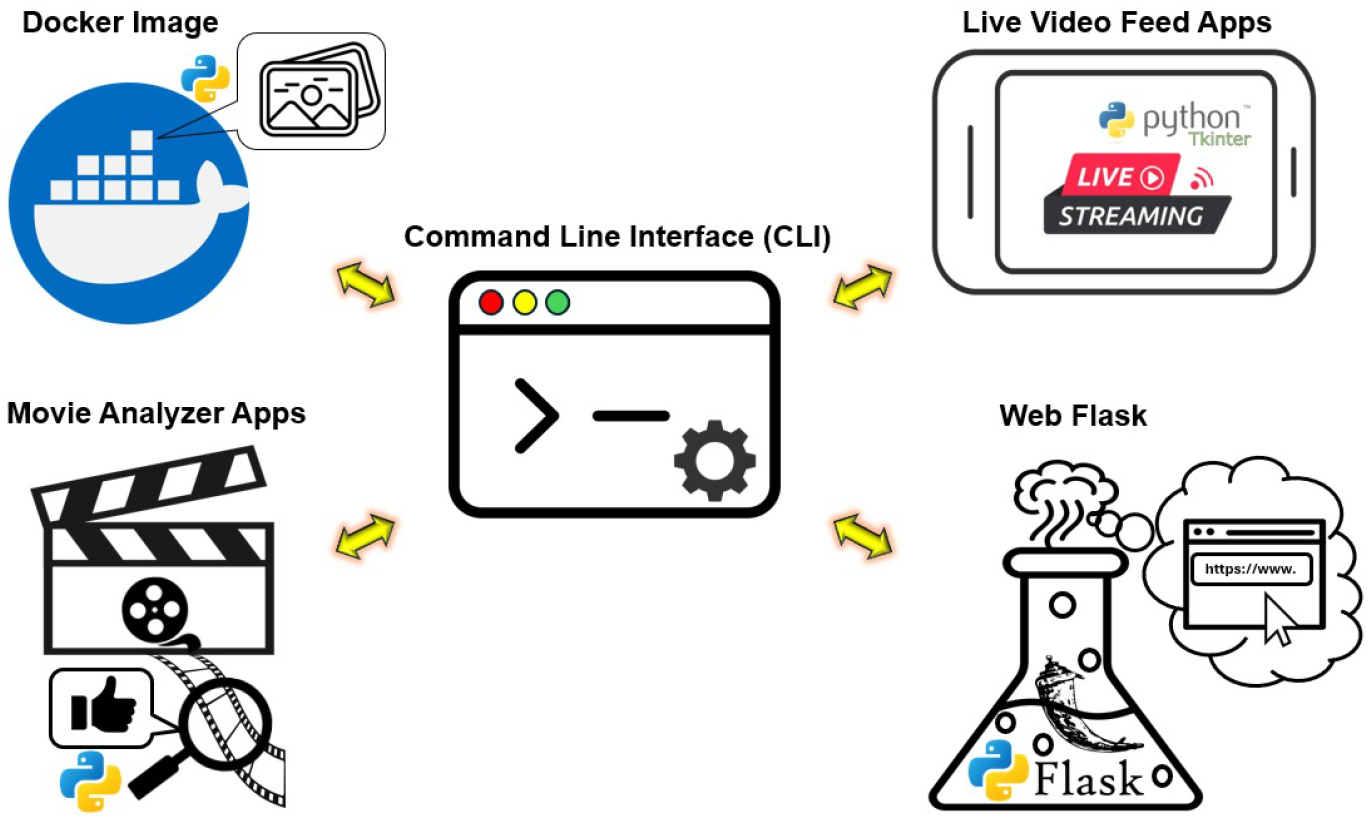
List of developed inference models using the trained AI model.

The overview of these inference models and brief description are shown in Table 1.

**Table 1:**
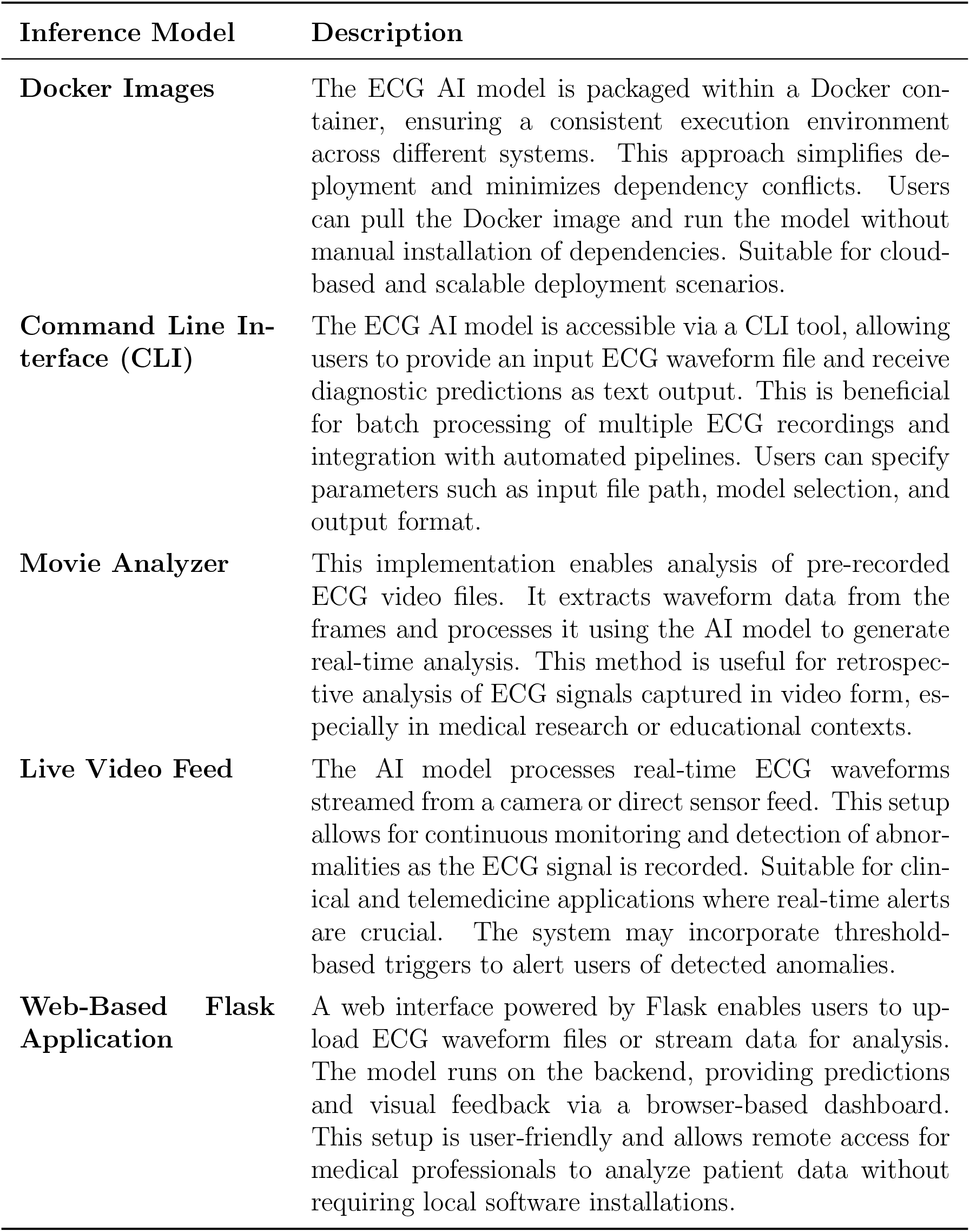
Overview of ECG Waveform AI Detection Inference Models.

Considering the docker image, the model can be executed and deployed in different systems to anonymize ECG scan images; this was achieved by including all the required dependencies and trained model into a tarball image. In the CLI model, the inference model was developed using Python programming language and automatically anonymizes multiple ECG scan images and crop out the waveform data from the uploaded scans using the terminal; this was tested extensively on GNU/Linux systems and requires Python interpreter and its libraries. The movie analyzer app can be used to perform waveform detection by analyzing a movie clip with mp4 format; this detects the waveform in each frame from the user supplied video clip. Similarly, the live video feed model utilizes gPhoto2 (http://www.gphoto.org/) to analyze the frames in the live video feed; this was tested on Nikon D800 digital single-lens reflex (DSLR) camera equipped with a 50 mm 1.8D Nikkor lens. Note that, to reduce the output file size and analysis time, some frames in the video analysis were set to be skipped. Lastly, a web-based flask application written in Python programming language was developed; this enables the users to execute and deploy the present model on their system and access the model interface using the web browser. The general algorithm for the developed inference model is shown in 1.

#### Algorithm 1

General algorithm of the inference model developed in the present work.

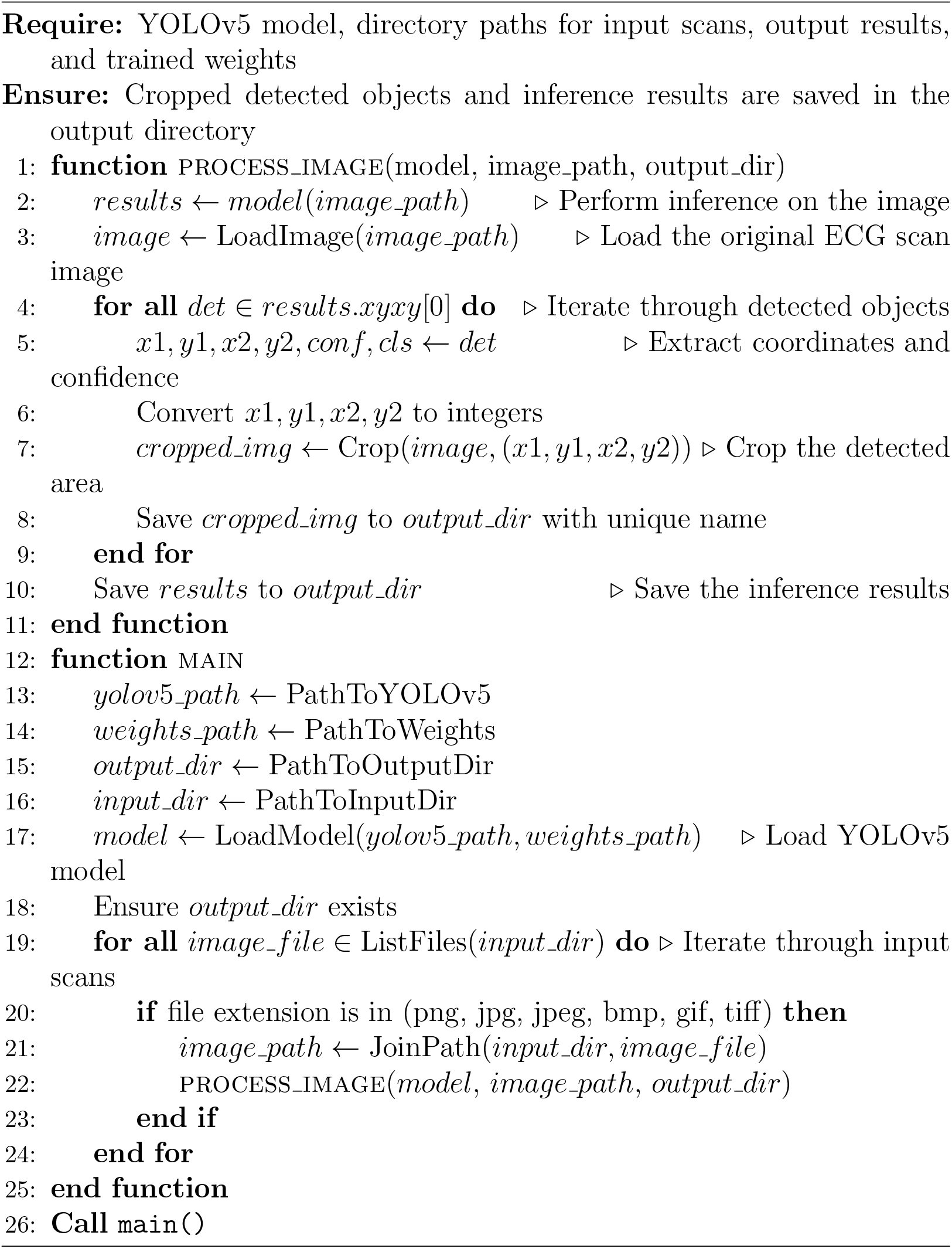

### 2.3 Validation scheme

In order to validate the present AI model, we have used number of different cases for each of the five inference models, namely, (1) scanned image of paper ECG, (2) Comparison between YOLO V5 and YOLO V9 version, (3) movie clip of multiple scanned images of paper ECG, (4) image of ECG scan taken from smartphone camera, (5) real-time detection of waveform from a printed ECG scan.

For the scanned ECGs, we selected six different ECG scans, each presenting diverse shadows, waveforms, and grid patterns.

YOLO (You Only Look Once) models are indeed among the top-performing approaches for real-time object detection. They balance speed and accuracy, making them widely adopted in applications where inference time is critical (e.g., robotics, autonomous driving, real-time surveillance). However, whether they are the absolute “state-of-the-art” depends on the specific task and evaluation metrics. In addition to training with YOLO V5, we also retrained our model using YOLO V9, a more recent version noted for its improved performance relative to YOLO V5. Therefore, we have re-trained our entire AI model based on a two-stage detector Fast R-CNN (FR-CNN) method that known to have improvement over the R-CNN by sharing convolutional layers. FR-CNN is the state-of-art method that combines classification and bounding box regression into a single network, improving both speed and accuracy. Its efficiency and simplicity laid the groundwork for subsequent advancements like Faster R-CNN and transformer-based detectors. All the developed models, source codes, and validation materials are freely available for download from: https://doi.org/10.6084/m9.figshare.26204303

For dynamic ECG detection, the movie clips of multiple ECG results were generated by screen recording while scrolling through these scans. These mp4 movie files were analyzed using our custom-developed movie analyzer app.

To evaluate the model performance in real-world application, we used images taken from smartphone cameras of ECG scans displayed on computer screens and paper prints, ensuring the robustness of our AI model against the various noises.

For the smartphone camera images, ECG scan taken from smartphone camera from a computer screen add realistic glare, shadow and orientation mismatch to the ECG scans that will be supplied to our AI model. We randomly selected 50 ECG scans and captured images of them using a smartphone camera. These scans were then divided into two identical groups: “ECG scans,” representing the original scans, and “ECG scans on screen,” referring to the images taken from a computer screen using the smartphone camera. Each group was separately analyzed by our model, utilizing the YOLO V5 version of our trained AI.

Real-time detection of waveforms from ECG paper prints was conducted using a live video feed model, particularly to assess the accuracy, speed, and detection capabilities of our AI model. In this case the waveform characteristics would be detected, selected and extracted by analyzing live feed from a DSLR camera.

All validations were performed on a GNU/Linux machine, and patient data was anonymized in all the presented results.

### 2.4 Evaluation of computational burden

In order to determine the central processing unit (CPU) load, we processed 495 ECG scans using our YOLO V5 version of our model. The analysis was conducted on a Meerkat computer from System76 (https://system76.com/), equipped with an 11th Gen Intel i7-1165G7 (8) @ 4.700GHz CPU and no dedicated GPU. The psutil library (https://pypi.org/project/psutil/) was utilized to monitor CPU load over time while running the model across all 495 ECG scans.

## 3 Results

### 3.1 Waveform detection in different shadows, waveforms, and grid patterns

The results of waveform detection for six different ECG results with diverse shadows, waveforms, and grid patterns are shown in Fig. 3. The detected waveform area is marked using a blue box with the confidence of inference shown above the box. The program automatically crops the selected area and saves the waveform in a separate directory. Considering the results shown in Fig. 3, our trained AI model could accurately detect, select, and extract the waveform data from ECG scans. Anonymizing user input ECG results and appropriately saving the waveform data are crucial steps for data standardization and integration into electronic health records. This process ensures the confidentiality and privacy of patient information while allowing the standardized data to be used effectively in medical records. Moreover, the anonymized output data generated from the current model can serve as a valuable resource for training other cardiac-related models, thereby enhancing the development and accuracy of these models without jeopardizing the privacy of patients. This approach not only promotes the secure handling of sensitive health information but also supports advancements in cardiac care and research through the use of high-quality, anonymized data.

**Figure 3.**
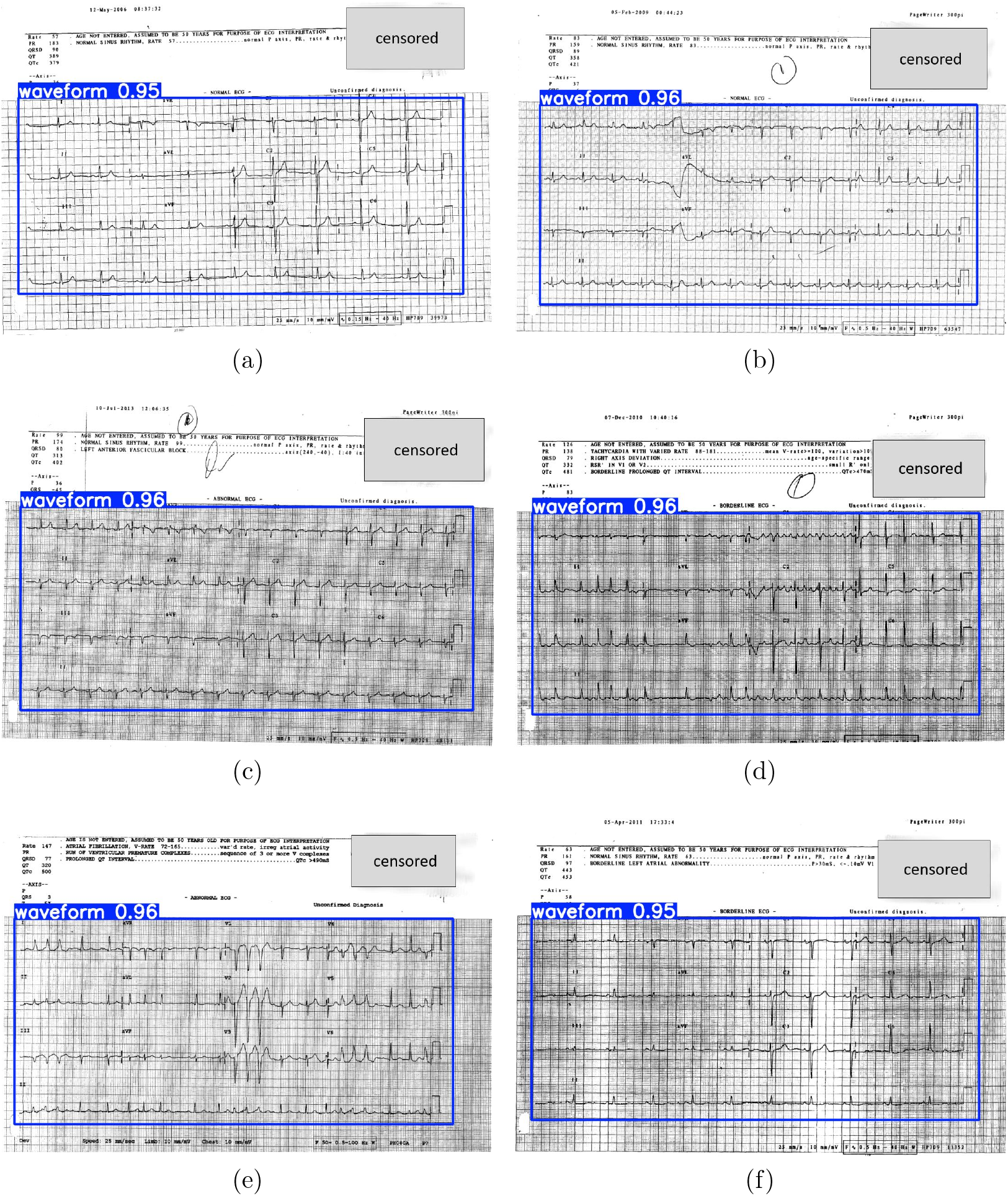
Waveform detection from six different ECG scans with diverse shadows, waveforms, and grid patterns.

### 3.2 YOLO V5 vs. YOLO V9

Fig. 4 compares the results of these two models. Based on this comparison, we concluded that both YOLO V5 and YOLO V9 perform adequately for waveform feature detection.

**Figure 4.**
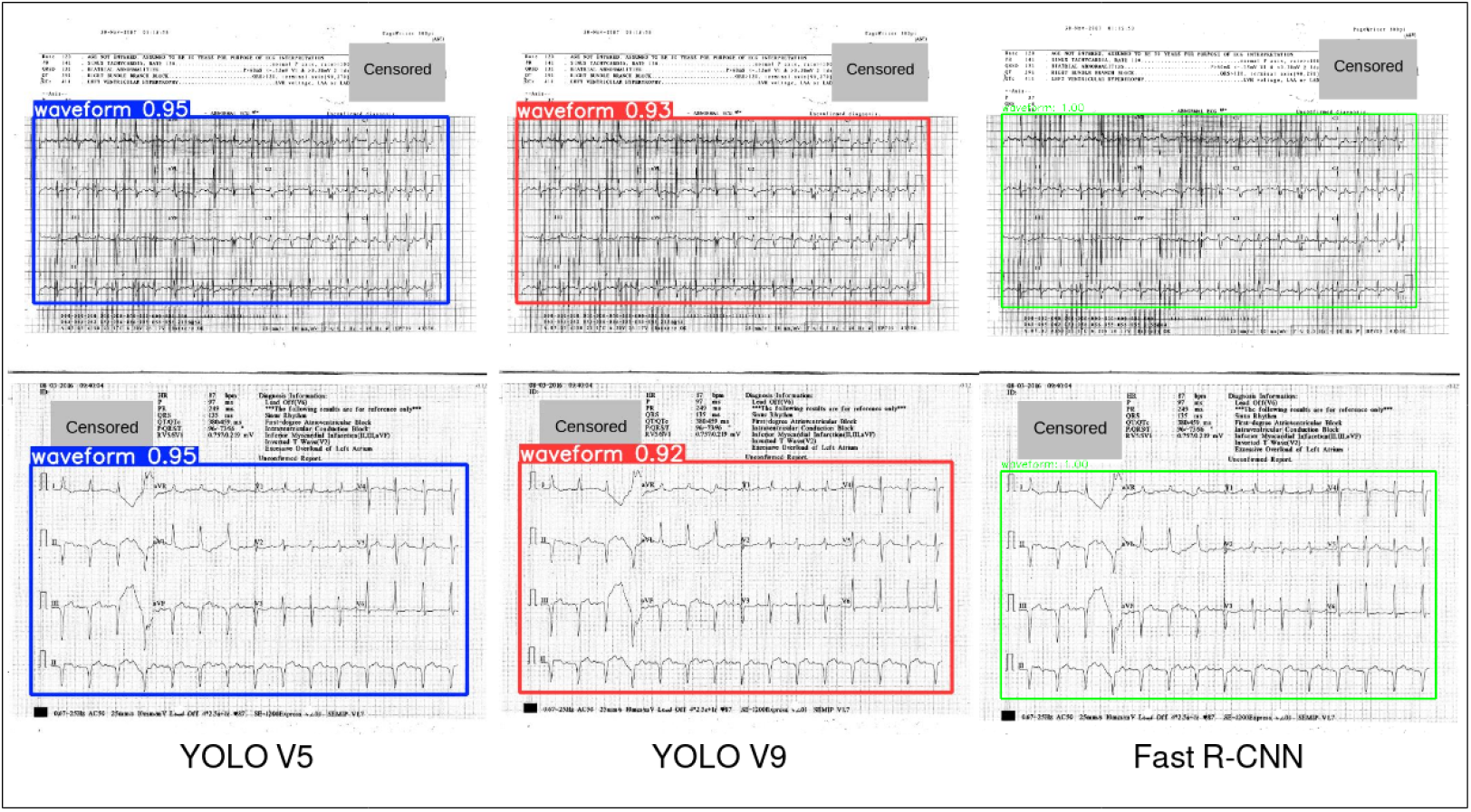
The Comparison between YOLO V5, YOLO V9 and Fast R-CNN trained models.

### 3.3 Dynamic ECG detection from videos

The analysis of a movie clip of multiple scanned images of paper ECG has been performed to assess the accuracy and robustness of the present AI model. Four frames of part of this analysis are shown in Fig. 5. The full video clip of the analysis can be viewed from: https://doi.org/10.6084/m9.figshare.26204303. Considering the analysis of this movie clip containing moving ECG scans, it can be concluded that our model could accurately detect and analyze moving waveform features at a high speed; this is particularly important when analyzing large volume of ECG data by bulk processing ECG scans. Through rapid and precise identification of waveform characteristics in motion, our model ensures that extensive datasets can be processed in a timely manner. This efficiency is critical for applications such as real-time monitoring, large-scale medical studies, and automated diagnostic systems, where the ability to rapidly and accurately analyze ECG data can lead to improved patient outcomes and more effective use of medical resources. The high-speed performance of our AI model in analyzing moving waveforms underscores its potential as a powerful tool in the field of cardiac care and research, facilitating the handling of large-scale ECG datasets with ease and precision.

**Figure 5.**
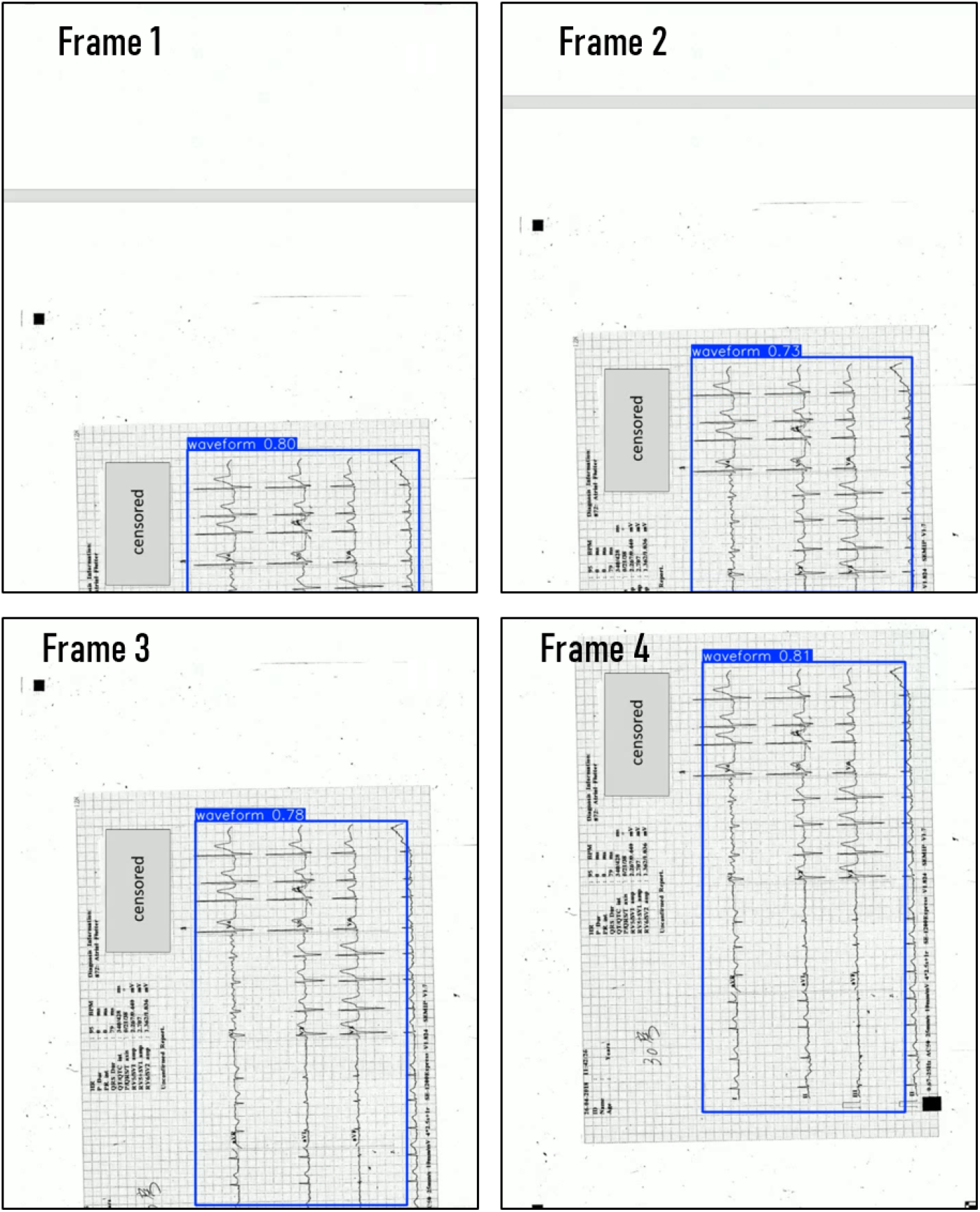
Four frames showing the waveform detection mechanism from a video clip of a scrolling ECG scan. The detection analysis can be viewed in the video clip at: https://doi.org/10.6084/m9.figshare.26204303

### 3.4 Real-world ECG from smartphone cameras

The snapshots and their analysis (i.e., waveform detection) are shown in Fig. 6. Considering substantial amount of glare and noise associated with the snapshots taken using smartphone camera from two different orientations, our model has the ability to satisfactorily detect, select and extract the waveform components from these images with a high confidence. The model effectively mitigates the challenges posed by varying lighting conditions and image quality, ensuring accurate extraction of the essential waveform data. This ability is crucial for processing images captured in less-than-ideal conditions, allowing for reliable analysis and interpretation of the waveform components. By leveraging advanced image processing techniques, our model enhances the precision and reliability of waveform extraction, making it a valuable tool for applications that require accurate data from smartphone-captured images. This feature will be particularly useful for future development of smartphone applications that can make use of our trained AI model to perform portable and real-time ECG feature detection and extraction.

**Figure 6.**
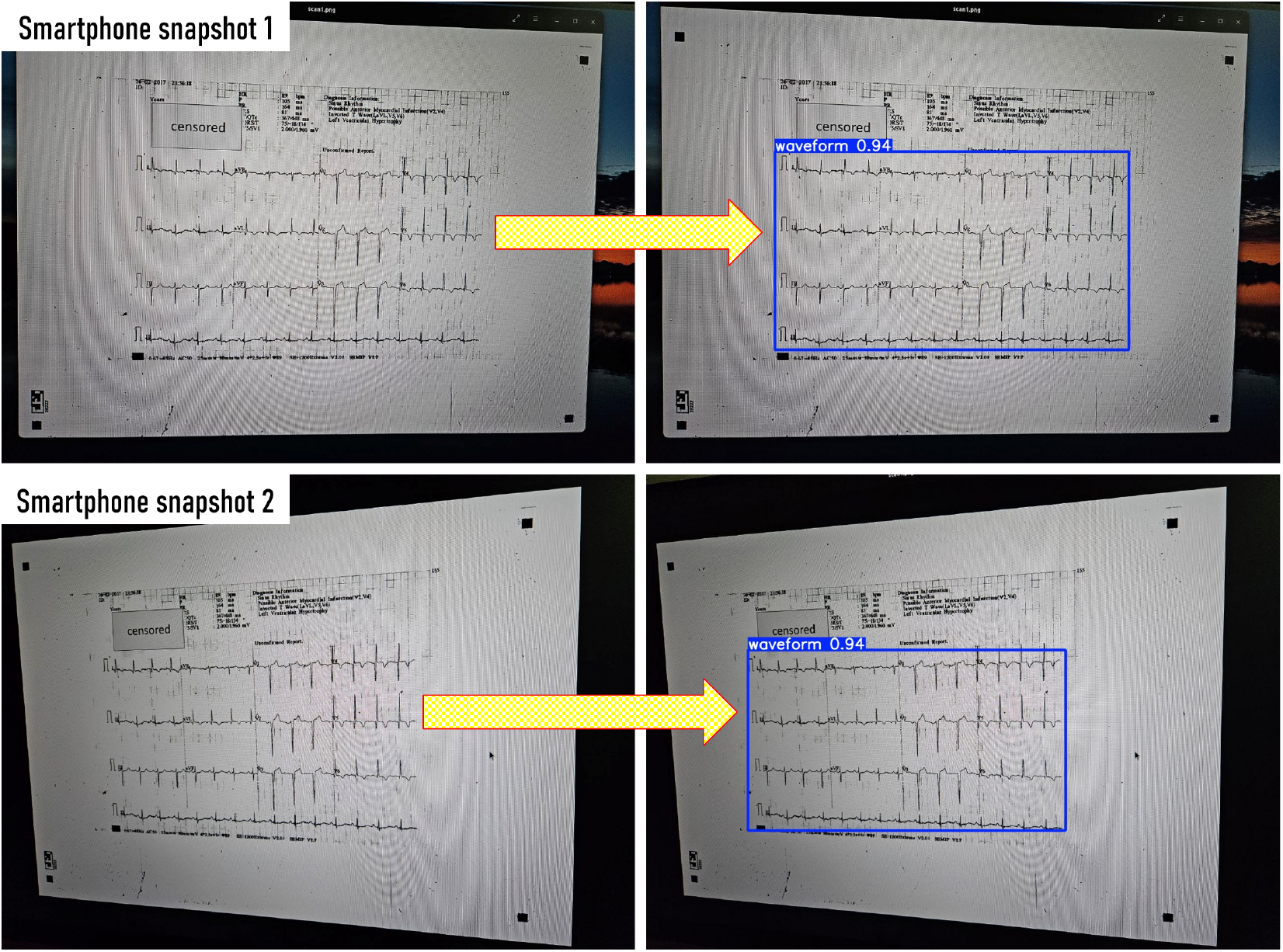
Waveform detection of images taken using smartphone camera from the computer screen at two different orientations.

We generated a histogram to visualize the confidence levels in detecting waveform features, as illustrated in the Fig. 7.

**Figure 7.**
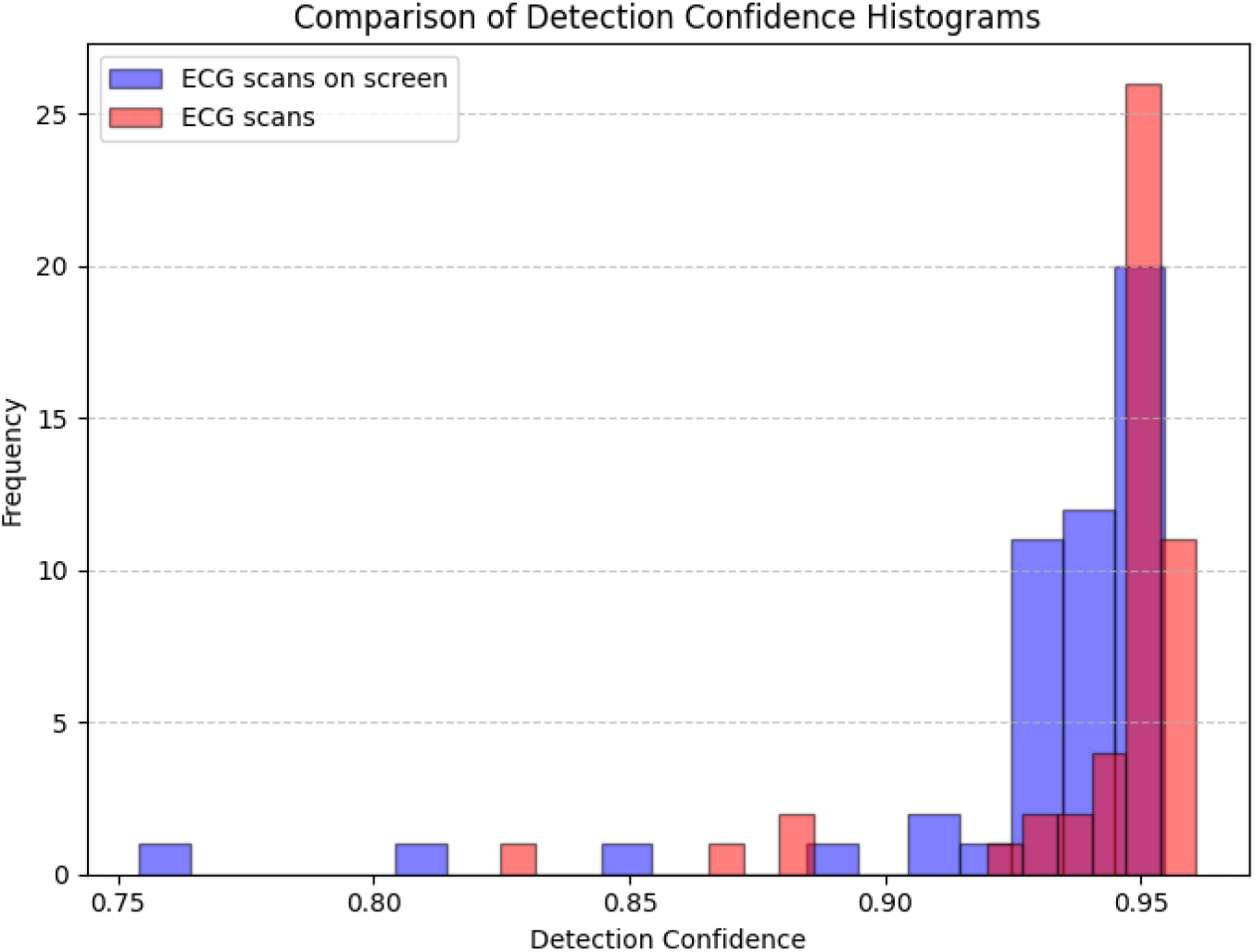
Comparison of detection confidence from ECG scans and ECG scans displayed on screen for 50 identical 12 lead ECGs.

### 3.5 Real-time ECG from printed scans

The results shown in Fig. 8 show six snapshots taken from the analyzed live video feed. The detection analysis of the live feed can be viewed in the video clip at: https://doi.org/10.6084/m9.figshare.26204303. Considering the results from this numerical experiment, we have noted that our model performs well in detecting waveform features from a live video feed captured through an external DSLR camera. The live feed detection method presents a novel application of our AI model that can be deployed in real-time health monitoring systems.

**Figure 8.**
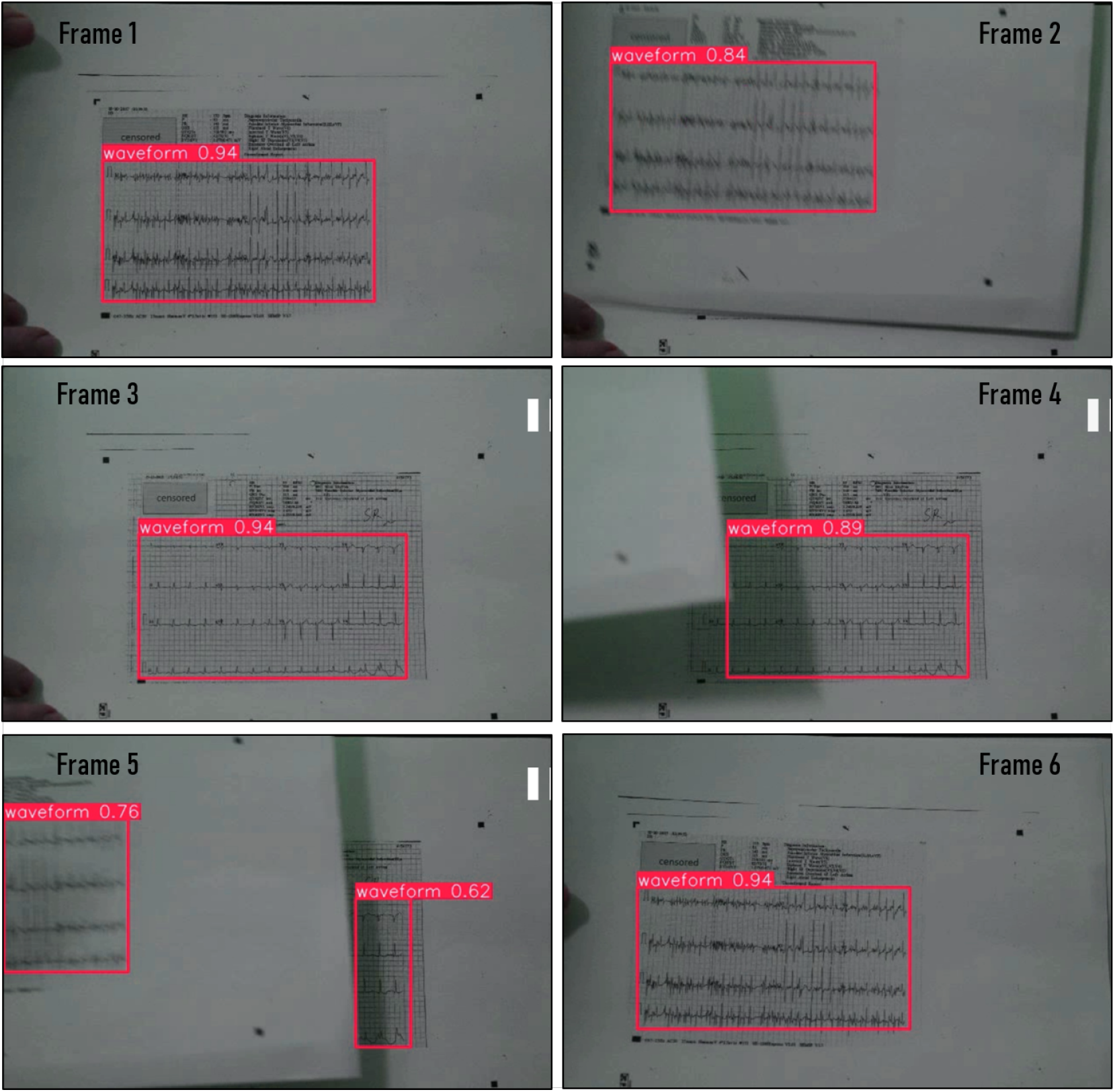
Six frames showing the waveform detection mechanism from a live video feed captured from a DSLR camera pointing at printed ECG scans. The detection analysis can be viewed in the video clip at: https://doi.org/10.6084/m9.figshare.26204303

A total of 495 ECG scans were employed, and for each detected waveform feature, comprehensive metrics—such as bounding box dimensions, detection area, and confidence scores—were extracted. In addition, the histograms of the number of detections per image and the distribution of confidence scores were generated and are presented in the Figs. 9 and 10.

**Figure 9.**
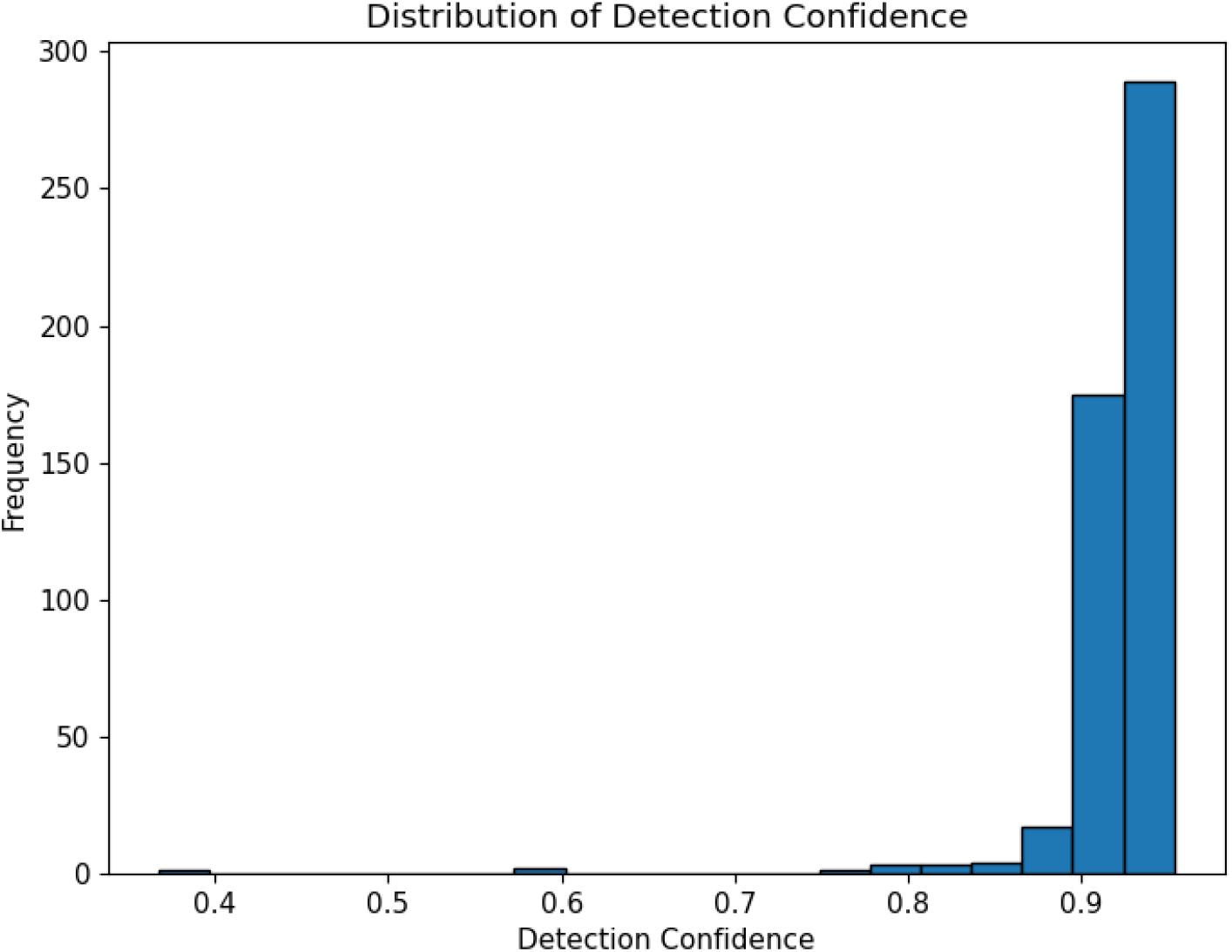
Detection confidence histogram.

**Figure 10.**
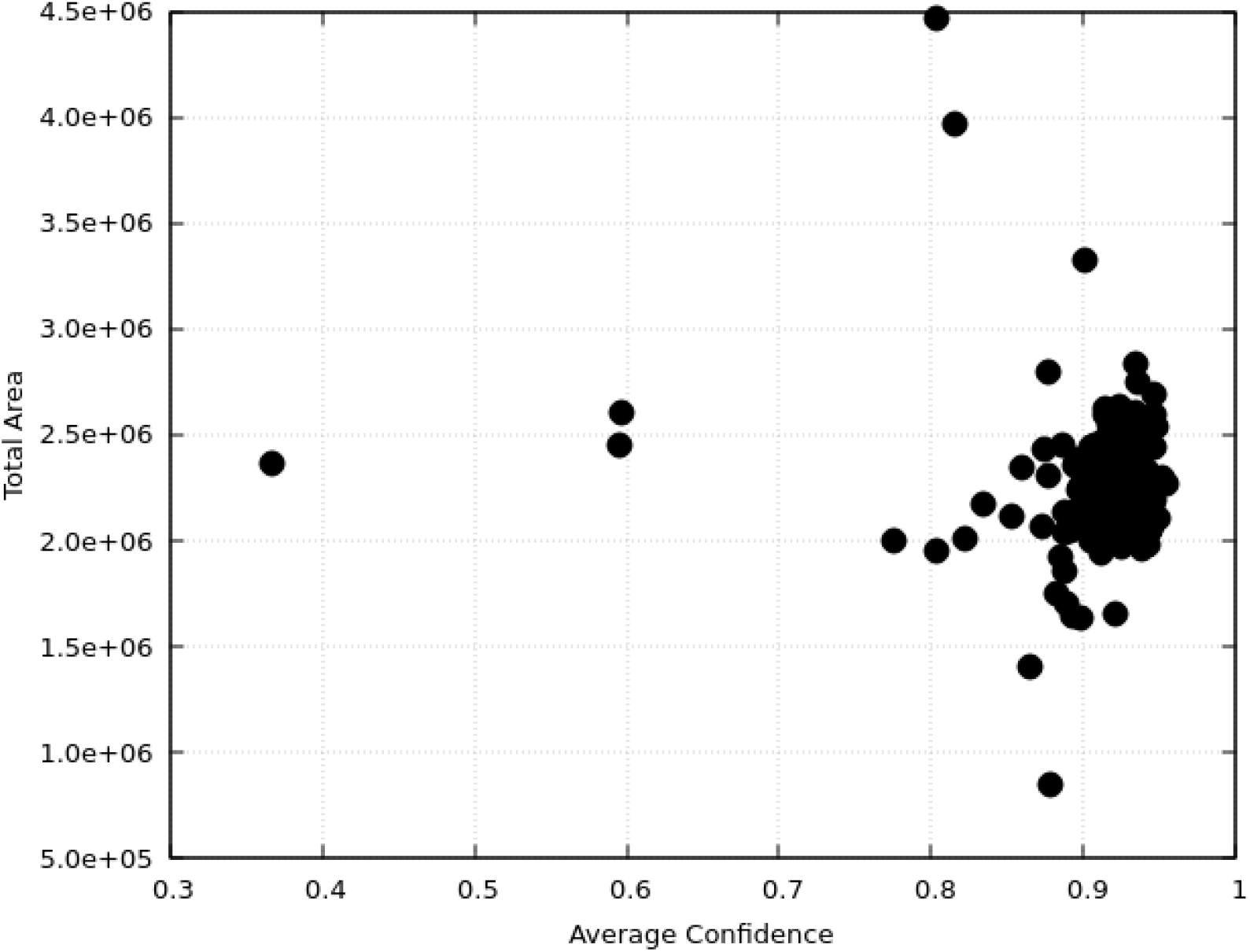
Average confidence versus total area of detected waveform feature.

### 3.6 Evaluation of computational burden

The results of CPU load are presented in the Fig. 11. The CPU usage was within 30% during the whole analysis process, ensuring efficient data processing without extra computational burden.

**Figure 11.**
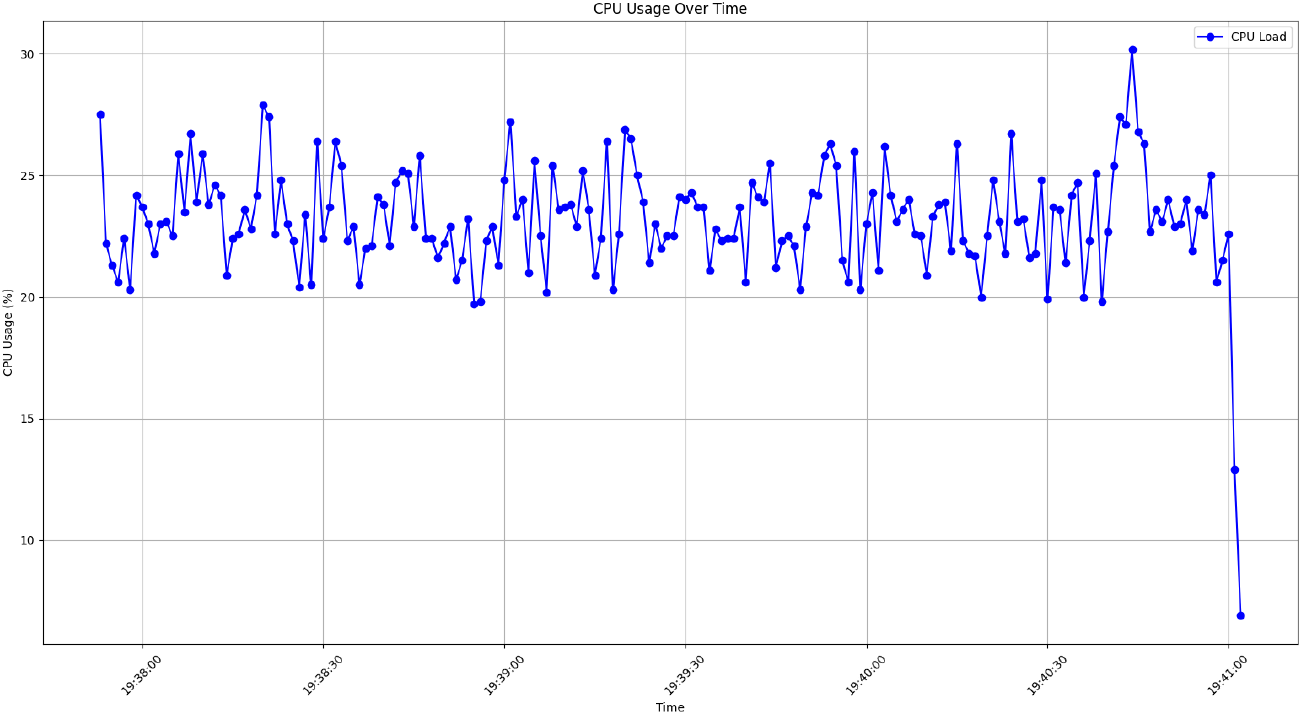
CPU load for analyzing large amount of ECG scans.

All the video clips, testing data, trained AI model and source codes were made openly and freely available under GPLv3 license and can be downloaded from: https://figshare.com/articles/software/Development_of_an_open-source_artificial_intelligence_AI_anonymizer_for_elecscans/26204303

## 4 Discussion

### 4.1 Summary of results

In the present work, we have developed an AI model that can detect, select and extract waveform features from ECG scans. Five tools were developed that are, namely, (1) docker images, (2) command line interface (CLI), (3) movie analyzer, (4) live video feed, (5) web-based flask application. The model was validated using four different schemes ranging from scanned ECG images, movie clips, ECG scans with glare and live video feed detection. The performance of the developed model and inference methods is adequate in detecting, selecting, and extracting waveform data in an accurate, high speed and robust fashion. Considering the results from four different numerical experiments that was performed in this validation scheme, These results suggest that the proposed AI model is robust, accurate and functional in detecting, selecting and extraction waveform features from variety of ECG scans. We believe the present model and development inference tools will be useful in automation of data curation, record keeping and analysis in cardiac related studies.

### 4.2 Unique advantages: comparison with existing works

Compared with prior ECG AI systems, our open-source anonymizer delivers real-time, multi-modal processing that localizes ECG waveforms first and then performs downstream tasks, whereas leading digitizers are optimized for scanned paper ECGs and can degrade with overlaps [5]. Synthetic dataset work similarly notes that digitization accuracy measured by correlation with ground truth-dropped below 60% in the presence of overlapping leads [15]. Our model is designed for varied inputs (smartphone photos, video frames, live feeds) where clutter and occlusion are common.

This complements but is distinct from diagnostic classifiers that target disease labels rather than waveform extraction. Methodologically, our detection first approach contrasts with earlier image only pipelines; prior work emphasizes two routes, namely the image of the ECG can be input directly into a 2D CNN architecture or a digitization pipeline to 1D signals [16].

Another notable unique advantage is integration of detection and anonymization in a single, open-source workflow that can anonymize only after high confidence waveform localization to preserve utility while minimizing false positives. This two stage design contrasts with end-to-end signal altering methods like PrivECG, a generative adversarial network (GAN) framework capable of privatizing 12-lead ECGs [17], and with commercial interpretation platforms focused on closed clinical outputs, e.g., PMcardio [https://www.powerfulmedical.com/]. By unifying detection/digitization and anonymization tasks typically handled separately, our model streamlines deployment and improves real-world applicability across diverse clinical and research settings.

### 4.3 Application scenarios

Real-world use case examples: (1) the live video feed from a patient’s wearable device can be sent to the healthcare provider. The our model automatically detects and crops the ECG waveform, preparing it for immediate analysis or archiving without needing additional equipment or software, (2) hospitals and clinics often need to share ECG data for research, training, or AI model development. This AI model can anonymize the ECG images or videos by automatically cropping out patient-sensitive information, such as names or IDs, ensuring privacy and compliance with data protection regulations, (3) many older ECG records exist as scans or videos of printed outputs.

The model can efficiently crop and digitize these ECG waveforms, excluding any handwritten or printed patient identifiers, making the data ready for archival, analysis, or integration into electronic health record (EHR) systems, (4) during emergencies, live ECG video feeds can include extraneous information such as patient details or irrelevant visual elements. The AI model can isolate the waveform in real-time, providing a focused and anonymized stream (to avoid any leak of patient’s private information) for analysis, remote consultations, or AI-based anomaly detection, (5) in telemedicine, patients might share ECG readings captured through phone cameras or low-cost devices. The model can detect and crop the waveform while removing sensitive data, ensuring secure sharing and maintaining data integrity for remote diagnosis, (6) medical educators often use ECG visuals for teaching purposes. The model can quickly extract waveforms from various sources, removing sensitive information, and creating clean visuals for presentations, tutorials, or online courses, (7) healthcare providers can use the model to crop and clean ECG visuals for use in patient reports or consultations, focusing only on the waveform and eliminating unnecessary or sensitive information to ensure clarity and privacy. Another real-world example that we have recently investigated was to employ our model to prepare clean and anonymized ECG waveforms for the ECG university at the International Society for Holter and Noninvasive Electrocardiology (https://ishne.org/ecg-university/), in which different well-known cardiologists submitted their cases studies to be used in the ECG university for educational and training purpose. Our developed AI model was then used to ensure all the waveforms are anonymized and without losing any important ECG waveform data.

### 4.4 Limitations and future directions

In future works, we aim to further expand the present model to detect individual ECG leads and digitize these leads to detect heart abnormalities using an AI powered system. The process of digitizing ECG waveforms presents several challenges that can impact data integrity and clinical interpretation. Errors such as signal clipping, particularly in the QRS complex, can distort critical features and lead to misclassification of arrhythmias or ischemia. Additionally, issues like inadequate sampling rates, baseline noise, lossy compression, and segmentation errors can further compromise waveform accuracy. Variability across different ECG devices and interpolation artifacts may also introduce inconsistencies. To mitigate these risks, applying robust signal processing techniques, ensuring standardized digitization protocols, and validating digitized waveforms against raw ECG recordings are essential steps in preserving data quality and clinical reliability. In future work, we will explore methods to enhance ECG digitization accuracy and minimize potential sources of error by expanding our model to digitize ECG scans.

## 5 Conclusions

In this work, we presented an open-source, detection-first AI system and toolchain, comprising Docker images, a CLI, a movie analyzer, a live-video pipeline, and a Flask web app, that accurately localizes ECG waveforms and extracts features across heterogeneous inputs. Validation on four settings (scanned images, movie clips, glare-affected scans, and live feeds) demonstrates robust, fast, and reliable performance in realistic, cluttered conditions. By coupling high confidence waveform localization with anonymization, the workflow enables privacy-preserving sharing and downstream analysis, complementing diagnostic classifiers and overcoming limitations of paper centric digitizers. These capabilities support automated curation, record keeping, education, and telemedicine, and can streamline research pipelines and EHR integration. Future work will extend the framework to ECG lead-level detection and digitization, and address known risks in waveform recovery (e.g., clipping, sampling constraints, baseline noise, compression, and segmentation errors) through standardized protocols and validation against raw ECG recordings. Overall, the proposed system advances practical, real world ECG extraction and anonymization, providing a foundation for scalable cardiac data workflows.

## Patient Consent

This study received Ethics Approval from The Joint Chinese University of Hong Kong – New Territories East Cluster Clinical Research Ethics Committee (CUHK-NTEC CREC 2019.338 and 2019.361). Institutional board approval was obtained and waived the need for patient consent in this retrospective study.

## Data Availability

All data produced in the present study are available upon reasonable request to the authors.

https://www.proquest.com/docview/3252768154/278E578A36174385PQ/2?sourcetype=Dissertations%20&%20Theses

## Acknowledgments

This study is supported by a Research Impact Fund by the Hong Kong Metropolitan University (RIF/2022/2.2). This study arose directly from clinical data collected as part of GT’s doctoral thesis at the Faculty of Medicine, Division of Graduate Studies, The Chinese University of Hong Kong accompanied by an ethics approval from the Joint NTEC-CUHK Clinical Research Ethics Committee (CUHK-NTEC CREC 2019.338 and 2019.361).

